# A 4-year review of amniocentesis at a tertiary hospital in Ethiopia: an experience from a LMIC

**DOI:** 10.1101/2023.11.10.23298380

**Authors:** Haile G. Mekuria, Hilkiah K. Suga, Gadise Bekele

**Affiliations:** Department of Obstetrics and Gynecology, Myungsung Medical College/MCM Comprehensive Specialized Hospital, Addis Ababa, Ethiopia; Department of Research, Blue Health Medical Consultancy, Addis Ababa, Ethiopia; Department of Biostatistics, Addis Continental Institute of Public Health, Addis Ababa, Ethiopia

**Author notes:** First author (Haile G. Mekuria).

**Keywords:** amniocentesis, advanced maternal age, prenatal genetic study, chromosomal abnormality, fetomaternal complication

## Abstract

**Background:** Amniocentesis is a technique for withdrawing amniotic fluid from the uterine cavity using a needle via a trans-abdominal approach. It has multiple diagnostic and therapeutic uses such as prenatal genetic studies and amnioreduction for polyhydramnios. The commonest indications are advanced maternal age and increased risk on maternal serum screening. The objective of this study is to review the common indications, outcomes and complications of amniocentesis at a tertiary hospital in Ethiopia.

**Methods:** A retrospective cross-sectional study was conducted at a tertiary hospital in Ethiopia. Data was collected from the Fetomaternal division’s log records and follow-up charts during the time period of 2019 to 2022. All records of patients who had amniocentesis done were included in the study. Socio-demographic information and obstetric history and variables related to the procedures were retrieved. Descriptive statistics was done using the software IBM Statistical Package for Social Sciences (SPSS) version 25.

**Results:** A total of 35 patients’ records were reviewed. The mean maternal age was 32 years (range, 19 – 43 years). The mean gestational age was 20.9 weeks (SD ± 2.2). The commonest indications for amniocentesis were previous history of Down syndrome (11%), advanced maternal age (9%) and QUAD 1:200 for Down syndrome (6%). Karyotype test was done in 29 of the 35 amniocentesis cases which was normal in 72% of the cases, 17% had Trisomy-21 and 11% had Trisomy-18. Fifty-two percent of the pregnancy had a live full term delivery and 25% of the cases terminated the pregnancy.

**Conclusion:** The most common indications for amniocentesis in Ethiopia were abnormal ultrasound finding, advanced maternal age and abnormal QUAD test results. Around a quarter of the genetic tests which were done after the procedure had chromosomal abnormalities, from which, the majority were terminated. There was no complication related to the procedure.

## Introduction

Amniocentesis is a technique for withdrawing amniotic fluid from the uterine cavity using a needle via a trans-abdominal approach. Amniocentesis has been widely used since it was first performed in 1956. Originally reported as a method of determining fetal sex in utero, Fuchs and Riis then hypothesized that it could be possible to diagnose chromosomal abnormalities in utero via this technique. By 1963, it had been confirmed that the karyotype of fetal and amniotic cells are identical (1 – 3).

The most common diagnostic indications for obtaining amniotic fluid are prenatal genetic studies and assessment of fetal lung maturity. Other indications include, but are not limited to, evaluation of the fetus for infection, degree of hemolytic anemia, blood or platelet type, hemoglobinopathy, and neural tube defects. Risks include spontaneous abortion, infection, needle injuries, and preterm labor (4, 5).

Amniocentesis is also performed as a therapeutic procedure to remove excess amniotic fluid, such as in symptomatic polyhydramnios or twin-twin transfusion syndrome, or to reduce volume and pressure of amniotic fluid in cases of prolapsed fetal membranes in the second trimester to facilitate placement of an emergency cerclage.(6, 7)

Since the late 1960s, amniocentesis has become a widely accepted method of obtaining fetal genetic information (8). Amniocentesis for prenatal genetic studies is technically possible at any gestational age after approximately 11 weeks of gestation, but is optimally performed after 15 to 17 weeks of gestation. Procedures performed before 15 weeks (ie, early amniocentesis) are associated with higher fetal loss and complication rates, including culture failure, and should be avoided. Later procedures can be problematic if termination of pregnancy is planned based upon abnormal results. Although some pain is associated with amniocentesis, it is generally well tolerated without the need for anesthesia (8, 9).

Maternal age over 35 is the most common indication for amniocentesis (10). It is conventionally performed between 16 – 24 weeks. The chances of late-onset abnormalities after 1st and 2nd-trimester ultrasound examinations are estimated at 5.5%-17% (11).

Counseling about the risk and benefit of the procedure is very important as some women may decline the procedure if they don’t want to terminate the pregnancy whatever the result is. Others, even if they don’t want to terminate the pregnancy, they accept the procedure to prepare themselves for the care of the baby after delivery (12).

Prenatal chromosomal microarray analysis is recommended for a patient with a fetus with one or more major structural abnormalities identified on ultrasonographic examination and who is undergoing invasive prenatal diagnosis (13).

Even though the procedure is more than 6 decades old, as far as the authors search goes, this report is the first of its kind from an experience located in the Eastern Africa. This study aims to review the common indications and outcomes of prenatal diagnostic amniocentesis and its complications at a tertiary center in Ethiopia.

## Methods

A retrospective cross-sectional study was conducted at the Fetomaternal division, Department of Obstetrics and Gynecology, MCM Comprehensive Specialized Hospital, Addis Ababa, Ethiopia. The department is one of the few in the country that gives prenatal amniocentesis service. Data was collected from the division’s log records and follow-up charts which was abstracted from 2019 to 2022. Study period was from June 1^st^ to 30^th^, 2023. Records of all patients who had prenatal amniocentesis done during the timeframe were included in the study. The following variables were collected: indication, maternal age, gestational age, needle size, BMI, pregnancy outcome, result of the genetic test, presence of vaginal bleeding post-procedure, presence of amnionitis. The procedures were carried out by the same fetomaternal specialist using the same ultrasound guided techniques. IBM Statistical Package for Social Sciences (SPSS) version 25 was used to compute descriptive statistics.

Ethical approval was obtained prior to the data collection and the investigators did not collect personal identification information of the patients.

## Results

A total of 35 women had amniocentesis between 2019 and 2022 with the mean maternal age was 32 years (range, 19 – 43 years). The mean gestational age was 20.9 weeks (SD ± 2.2, range 17 – 25) and mean maternal BMI was 21 kg/m2 (SD ± 2.4). In 23 patients, 20 Gauge needle was used and 22 Gauge was used in the others. The needle insertion to the probe angle was perpendicular in 30 of the procedures (85%) and parallel in 5 of the procedures (15%).

The commonest indications were previous history of down syndrome (11%), advanced maternal age (9%), cleft lip (6%), history of Recurrent pregnancy loss or RPL (6%), hydrocephalus (6%), non-immune hydrops (6%), omphalocele (6%), QUAD 1:200 for down syndrome (6%).

Karyotype test was done in 29 of the 35 amniocentesis cases. Karyotype results were normal in 72% of cases (21/29), Trisomy-21 in 17% (5/29) and Trisomy-18 in 11% (3/29). Almost half (52%) of the cases had a live birth, 46% (16/35) had a term delivery and 6% (2/35) had a preterm delivery. Stillbirth or miscarriage occurred in 17% (6/35), 25% (9/35) of the cases terminated the pregnancy and the outcome was not known in 6% (2/35) of the cases. Pathological ultrasonography has the highest positive detection rate (30%)

## Discussion

Amniocentesis is generally done after the 15th gestational week with a miscarriage risk of less than 1% (14, 15). The risk of spontaneous fetal losses following amniocentesis preformed before this gestational age increases (16). Likewise, the patients in our study were between the gestational age of 17 and 25 weeks, which were under the accepted gestational age range.

Currently twenty and twenty-two gauge needles are commonly used and in this study, two-third of the procedure was done using a 20G needle. The larger needle size i.e. 20G is associated with significantly shorter duration of amniocentesis. However, there is no significant difference in the intrauterine bleeding at the insertion site and fetal loss (17, 18). Likewise, none of the patients in this study had documented intrauterine bleeding in both groups (20G vs 22G).

A study done in South East China that did a retrospective analysis on more than 4700 patients who had an amniocentesis showed the commonest indications for amniocentesis were increased risk on maternal serum screenings and advanced maternal age. They are indications for 86% of the procedures (5). Another 5 year reviews from Malaysia and China hospitals showed similar findings i.e. both indications accounted for more than half of the total procedures (19, 20). Other common indications include poor obstetric history, pathological ultrasonography findings and positive results from non-invasive prenatal testing. However, in our study, the majority of indication for amniocentesis was done due to the pathological ultrasonography findings (57% vs 12% or 4%) followed by previous Down syndrome baby and increased risk on maternal serum screenings, each accounted for 11% of the procedures. Advanced maternal age was an indication in only 9% of cases.

Positive detection rate of pathological ultrasonography for chromosomal abnormalities is less than 15% (19, 20). Noninvasive prenatal DNA testing has the highest positive detection rate with greater than 40% (42% and 53%). In our study, pathological ultrasonography has the highest positive detection rate (30%).

## Conclusion

A review of the first 5 years of amniocentesis at our center showed that the most common indication to the procedure was abnormal ultrasound finding. Advanced maternal age and abnormal QUAD test results were also among the common indications. Around a quarter of the genetic tests which were done after the procedure had chromosomal abnormalities, from which, the majority were terminated. There was no complication related to the procedure. Further study is required to understand more fetomatenal complications of amniocentesis in Ethiopia.

## Data Availability

The data underlying the results presented in the study are available from the MCM Comprehensive Specialized Hospital data access.

---

**Table 1:**
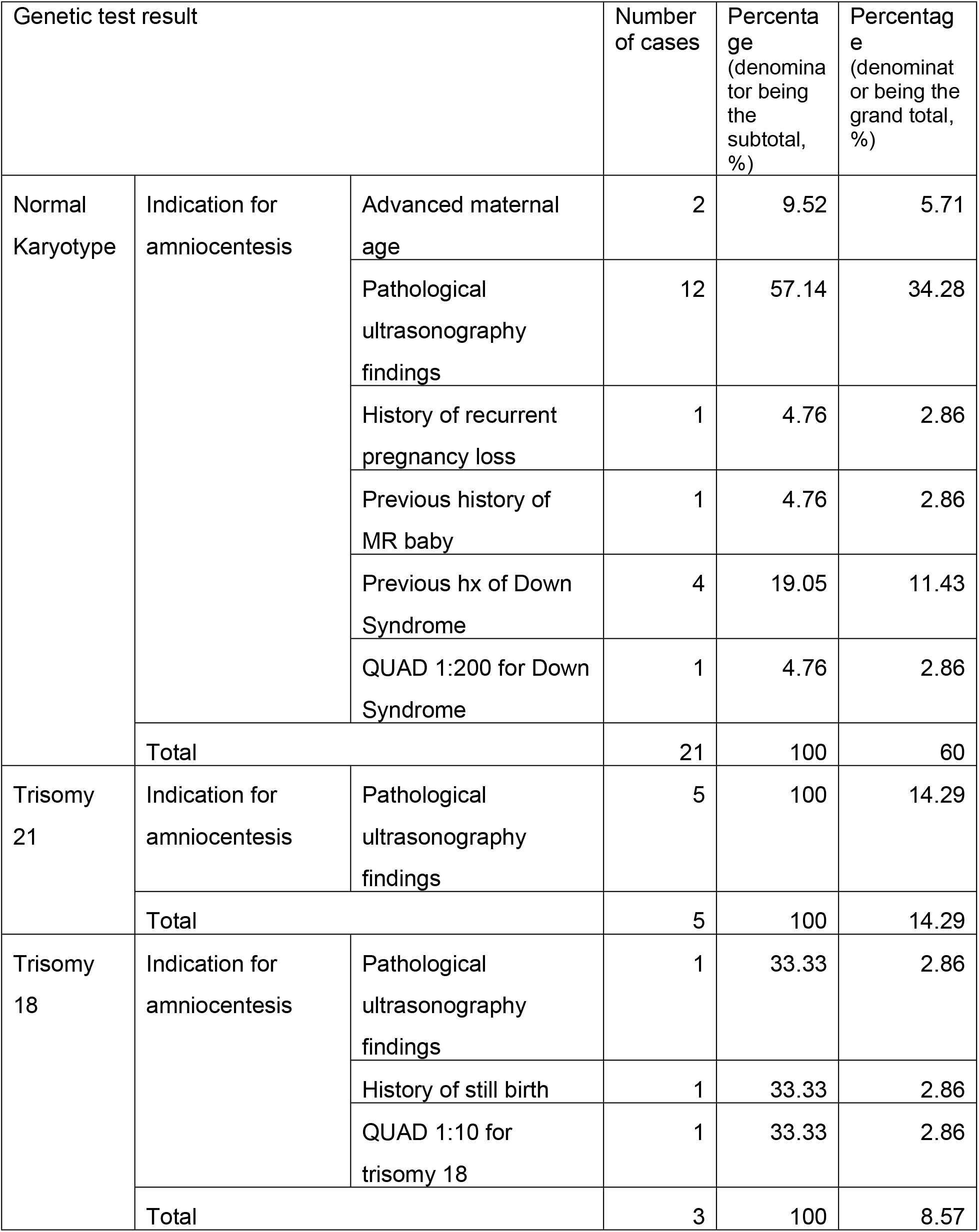
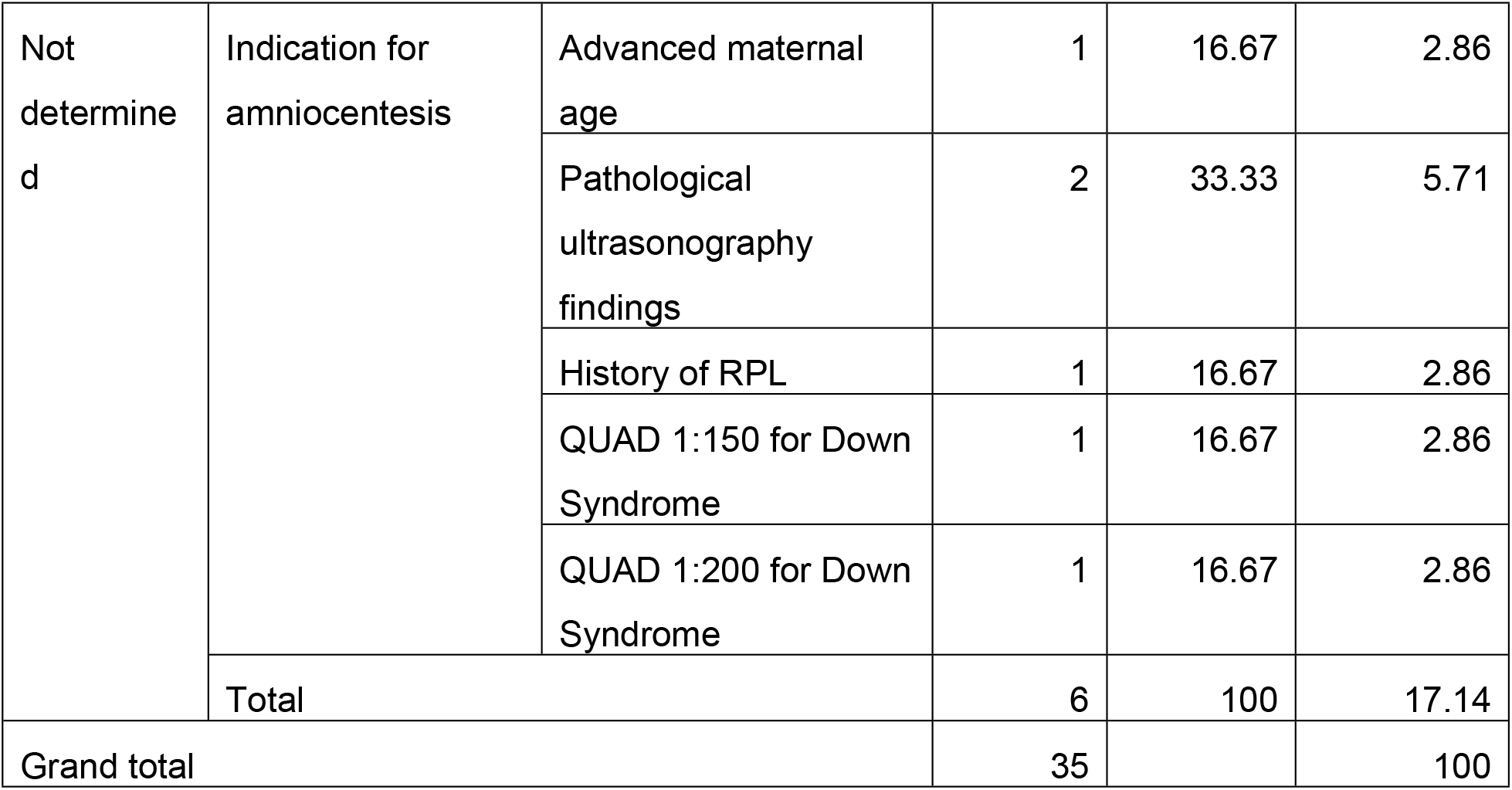
A five year summary of genetic test results and indications for amniocentesis at MCM Comprehensive Specialized Hospital, 2019 – 2022.

**Table 2:** Pregnancy outcomes and genetic test results of women who had amniocentesis at MCM Comprehensive Specialized Hospital, 2019 – 2022.

|  |  | Genetic test result |  |  | Total |
| --- | --- | --- | --- | --- | --- |
|  |  | Normal | Trisomy<br>18 or 21 | Not<br>determined |  |
| Pregnancy<br>outcome | Term delivery | 12 | 0 | 4 | 16 |
|  | Preterm delivery | 2 | 0 | 0 | 2 |
|  | Terminated | 1 | 7 | 1 | 9 |
|  | Still birth | 5 | 1 | 0 | 6 |
|  | Unknown | 1 | 0 | 1 | 2 |
| Total |  | 21 | 8 | 6 | 35 |

